# Clinical and genetic risk factors and long-term outcomes of MRI vessel wall enhancement in moyamoya disease

**DOI:** 10.1101/2023.05.20.23290282

**Authors:** Fangbin Hao, Cong Han, Mingming Lu, Yue Wang, Gan Gao, Qiannan Wang, Shitong Liu, Simeng Liu, MinJie Wang, Rimiao Yang, Zhengxing Zou, Dan Yu, Caihong Sun, Qian Zhang, Houdi Zhang, Qing-Bao Guo, Xiaopeng Wang, XuXuan Shen, Heguan Fu, JingJie Li, Bin Ren, Hui Wang, Hongtao Zhang, Huaiyu Tong, Wanyang Liu, Zhenghui Sun, Jianming Cai, Lian Duan

**Author notes:** Corresponding Author: Wanyang Liu, No.77 Puhe Road, Shenyang North New Area, Shenyang 110122, Liaoning Province, China; Zhenghui Sun, No.28 Fuxing Road, Haidian District, Beijing, 100853, China. Jian-ming Cai, No.8 Dong-Da Street, Fengtai District, Beijing 100071, China. Lian Duan, No.8 Dong-Da Street, Fengtai District, Beijing 100071, China. These authors contributed equally to this work (Fangbin Hao, Cong Han, Mingming Lu, Yue Wang).

## Abstract

**Background:** Intracranial vessel wall enhancement (VWE) on high-resolution magnetic resonance imaging (HRMRI) is associated with the progression and poor prognosis of moyamoya disease (MMD). However, the genetic background and risk factors for VWE in MMD have not been investigated. Therefore, this study assessed potential risk factors for VWE in MMD.

**Methods:** We evaluated MMD patients using HRMRI and traditional angiography examinations. The participants were divided into VWE group and non-VWE group based on the presence or absence of VWE on HRMRI. Logistic regression was performed to compare the risk factors for VWE in MMD. The incidence of cerebrovascular events of the different subgroups according to risk factors were compared using Kaplan – Meier survival and Cox regression.. STROBE (Strengthening the Reporting of Observational Studies in Epidemiology) guidelines were followed for reporting our cohort study.

**Results:** We included 283 MMD patients, 84 of whom had VWE on HRMRI. The VWE group had higher modified Rankin Scale scores at admission (*P* = 0.014) and a higher incidence of ischemia and hemorrhage (*P* =0.002) than did the non-VWE group. Multiple logistics regression shows risk factors for VWE included the ring finger protein 213 (*RNF213*) pR4810K variant (OR: 2.01 [95% confidence interval (CI), 1.08 - 3.76]; *P* = 0.028), hyperhomocysteinemia (HHcy; OR: 5.08 [95% CI, 2.34 - 11.05], *P* < 0.001), and smoking history (OR, 3.49 [95% CI, 1.08-11.31], *P* = 0.037). During the follow-up of 63.9 ± 13.2 months (medium 65 months), 18 recurrent stroke events occurred. Cox regression showed that VWE and *RNF213 pR4810K* variant were risk factors for follow-up stroke.

**Conclusion:** The *RNF213 p.R4810K* variant is strongly associated with VWE and poor prognosis in MMD. HHcy and smoking are independent risk factors for VWE; both are likely to induce the VWE phenomenon by affecting vascular endothelial function or causing vascular endothelial damage.

## Introduction

Moyamoya disease (MMD) affects the end part of the internal carotid artery (ICA) or the beginning part of the proximal branches, causing progressive stenosis and eventual occlusion.^1, 2^ Obtaining stenosed vascular tissue is challenging due to the location of the lesion; thus, the etiology of MMD remains unknown. Many imaging modalities are available for visualizing the affected blood vessels, including digital subtraction angiography (DSA), computed tomography angiography (CTA), and magnetic resonance angiography (MRA).^2^ However, these techniques cannot differentiate the vascular pathologies of MMD because different pathologies may result in the same luminal defects. Pathological lesions in MMD usually occur within the vessel wall. High-resolution magnetic resonance imaging (HRMRI) can be used to observe the vessel wall directly, potentially improving the visualization of pathological lesions in MMD.^3^ Vessel wall enhancement (VWE) is a unique HRMRI sign associated with stroke and MMD progression.^4^ The vessel wall morphology and its enhanced appearance provide additional information unobtainable from conventional luminal imaging methods.

The etiology of MMD is strongly linked to genetic factors.^5^ Ring finger protein 213 (*RNF213*) was first identified as a pathogenic susceptibility gene in Japanese familial MMD.^6, 7^ Furthermore, a previous study proved that the *pR4810K* variant (*rs112735431*) in *RNF213* has been found to be significantly associated with early-onset and rapid progression in MMD.^8^ Therefore, we hypothesized that pathophysiological processes of MMD are strongly influenced by genetic factors. Different genotypes may lead to different pathological changes in the vessel wall structure, and this alteration can be observed on HRMRI, such as VWE. However, associations between genetic risk factors and VWE in MMD are still unknown, and whether other factors have influenced the emergence of VWE is also unknown. Accordingly, we integrated highly relevant single nucleotide polymorphisms (SNPs) in *RNF213* and vascular risk factors of MMD into our analyses to identify the genetic background and risk factors for VWE. Furthermore, our research aimed to clarify further the role of VWE in predicting stroke risk and prognosis in MMD, which may provide clinical guidance.

## Methods

### Study population

The supporting data for the findings of this study can be obtained from the corresponding author upon reasonable request. This study was approved by the Research Ethics Committee of the Fifth Medical Center of the PLA General Hospital (reference number: 20150622). All participants or their representatives provided written informed consent before participating. The patients were enrolled consecutively in the Department of Neurosurgery, Fifth Medical Centre, PLA General Hospital, Beijing., China from January 2016 to December 2018. All patients underwent DSA and met the 2012 diagnostic criteria of MMD.^9^ Patients who provided consent for HRMRI and genetic testing were eligible. Those with an incomplete clinical profile or a history of Down syndrome, brain tumors, atherosclerosis, vasculitis, neurofibromatosis type 1, cranial irradiation, meningitis, or other diseases possibly misdiagnosed as moyamoya syndrome (MMS) were excluded. The STROBE (Strengthening the Reporting of Observational Studies in Epidemiology) guidelines for cohort studies were followed to report all findings (Supplemental Material).^10^

Clinical data, including sex, age, family history, clinical manifestations, neurological status, and history of hypertension, diabetes, hyperlipidemia, hyperhomocysteinemia (HHcy), smoking, and alcohol consumption, were collected. The patients’ neurological status was evaluated using the modified Rankin Scale (mRS) score on admission, classified as good (≤2 points) or poor (≥3 points).^11^ Clinical manifestations were categorized as a transient ischemic attack, infarction, hemorrhage, or other. Hypertension was diagnosed if the systolic blood pressure was >140 mmHg or the diastolic blood pressure was >90 mmHg. The presence of hyperlipidemia was considered when the levels of low-density lipoprotein, total cholesterol, or triglycerides exceeded 1.58 mmol/L, 2.26 mmol/L, or 1.69 mmol/L, respectively. The presence of diabetes mellitus is determined by fasting blood sugar levels exceeding 126 mg/dL, 2-hour oral glucose tolerance test results exceeding 200 mg/dL, or a glycosylated hemoglobin level of 6.5% or higher. HHcy was considered present when the homocysteinemia (Hcy) exceeded 15 mmol/L. Alcohol intake was considered a factor at >40 g/day, and smoking when there was a ≥6-month history of smoking.^12^

### Imaging protocols

Two experienced neuroradiologists (Lu and Zhang) blinded to the grouping and clinical information reviewed all angiographic and HRMRI images. A third reader evaluated radiographic presentations in the event of a disagreement.

Radiographic presentations included the Suzuki stage and posterior cerebral artery (PCA) involvement, and a higher Suzuki stage was used when different sides were present.

We used previously reported^13^ imaging sequences to acquire conventional magnetic resonance imaging (MRI) and HRMRI data using a 3T MR scanner (MAGNETOM SKYRA, Siemens Healthcare AG) with an 8-channel head coil.

The presence of VWE of steno-occlusive stenosis or occlusion lesions of the carotid T-junction in MMD was determined based on its signal intensity on post-contrast images compared to corresponding pre-contrast images in Siemens magnetic resonance workstation (Siemens Syngo AV1.1; Siemens Healthcare AG, Erlangen, Germany). Patients with comparable pre-and post-contract vessel wall signal intensities were classified into the non-VWE group (Figure 1). Patients were assigned to the VWE group if their post-contrast images had a higher vessel wall signal intensity than the pre-contrast images (Figure 2).

**Figure 1.**
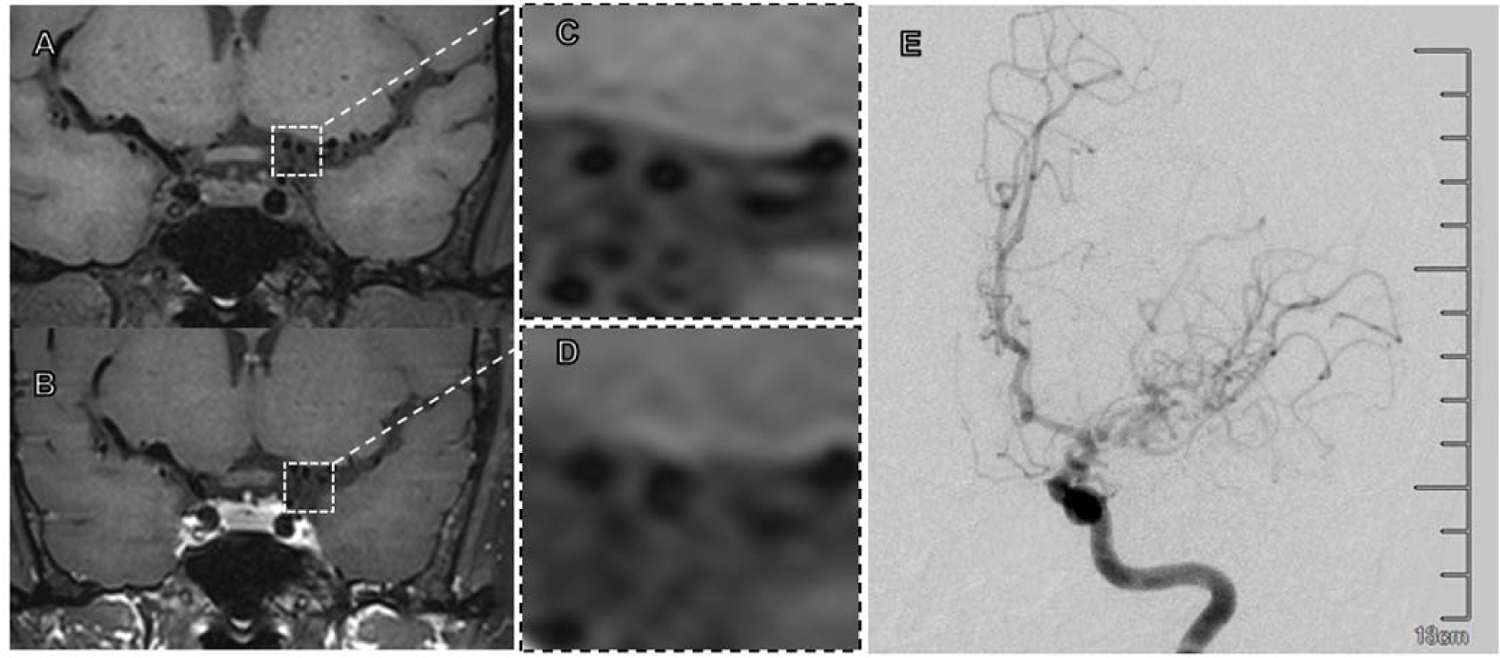
Representative high-resolution magnetic resonance images of a patient with moyamoya disease without vessel wall enhancement (VWE). (A, C) Pre-and (B, D) post-enhanced images show left distal internal carotid artery (ICA) stenosis without VWE (white square). (E) Stenosis of the left ICA and its branches on digital subtraction angiography.

**Figure 2.**
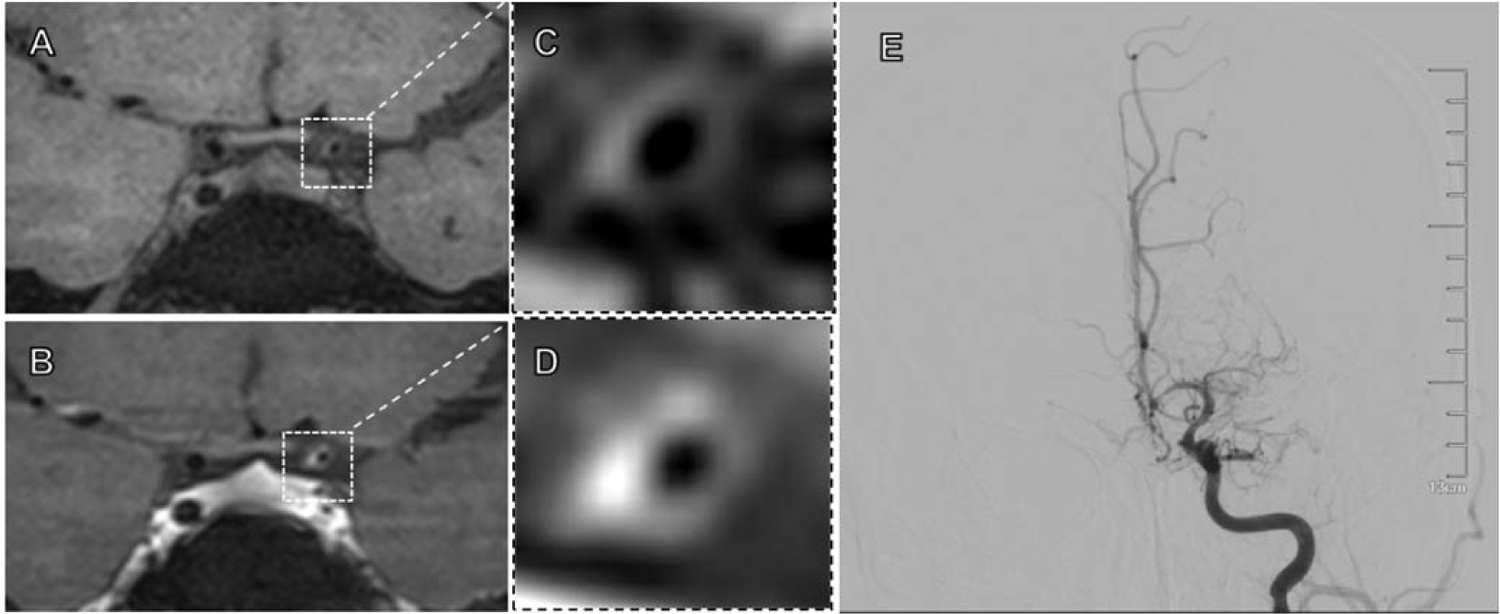
Representative high-resolution magnetic resonance images of a patient with moyamoya disease and vessel wall enhancement (VWE). (A, C) Pre-and (B, D) post-enhanced images show stenosis with a significant eccentric enhancement of the vessel wall in the left distal internal carotid artery (white square). The VWE degree is similar to that of the pituitary infundibulum. (E) Stenosis of the left carotid T on digital subtraction angiography.

### SNP selection and genotyping

Based on previous studies, *RNF213* p.R4810K (rs112735431) was selected owing to its association with early onset, rapid progression, and endothelial damage in MMD.^5, 8, 14–16^ Peripheral blood samples were collected at first admission. In addition, genomic DNA was extracted as previously described^17, 18^. The genotype of each sample was confirmed using fluorescent readouts.

### Follow up

Clinical outcomes were assessed through clinical visits or telephone interviews three to six months after discharge, with an annual follow-up thereafter. The medical charts were completed by researchers blinded to the patient’s genotype. During the follow-up period, stroke events were the primary endpoint of this study.. Ischemic and hemorrhagic strokes during the follow-up period were confirmed by CT or MRI.

### Statistical analyses

All statistical analyses were conducted using the R statistical software (www.R-project.org, The R Foundation) and Free Statistics software version 1.7^19^.

Normally and non-normally distributed continuous variables were compared using t-tests and rank-sum tests, respectively. Categorical variables were compared using the χ^2^ test or χ^2^ with Fisher’s exact test when appropriate. Univariate and multivariate logistic regression analyses were used to evaluate the effects of various clinical variables on the patient’s neurological status. Variables with *P*-values of < 0.05 in the univariate analysis were included in the multivariate analysis. Cox regression analysis and Kaplan-Meier (K-M) survival analysis were used to estimate the cumulative event-free rates based on the grouping of different risk factors. All tests were two-sided, and a *P*-value of <0.05 was considered statistically significant.

## Results

### Clinical and genetic differences between VWE and non-VWE group

In this study, 283 MMD patients were enrolled (Figure 3). The median onset age was 38.6 ± 9.9 years, the female-to-male ratio was 1.3:1.0, and 16 patients (5.7%) had a family history of MMD. There were 41.7% of patients with a transient ischemic attack (TIA), followed by 29.3% with a cerebral infarction, and 14.1% with a hemorrhage.. Eighty-four (29.7%) patients presented with VWE, and 73 (25.8%) had PCA involvement (Table 1). Sex, family history, Suzuki stage, PCA involvement, hypertension, diabetes, and hyperlipidemia did not differ between the VWE and non-VWE groups. However, the VWE group had a significantly lower onset age (P = 0.017), higher mRS score at admission (*P* = 0.014), and more incidences of cerebral infarction and hemorrhage (*P* = 0.002) than did the non-VWE group. Furthermore, significantly more patients had HHcy and a smoking history in the VWE group than in the non-VWE group (Table 1).

**Figure 3.**
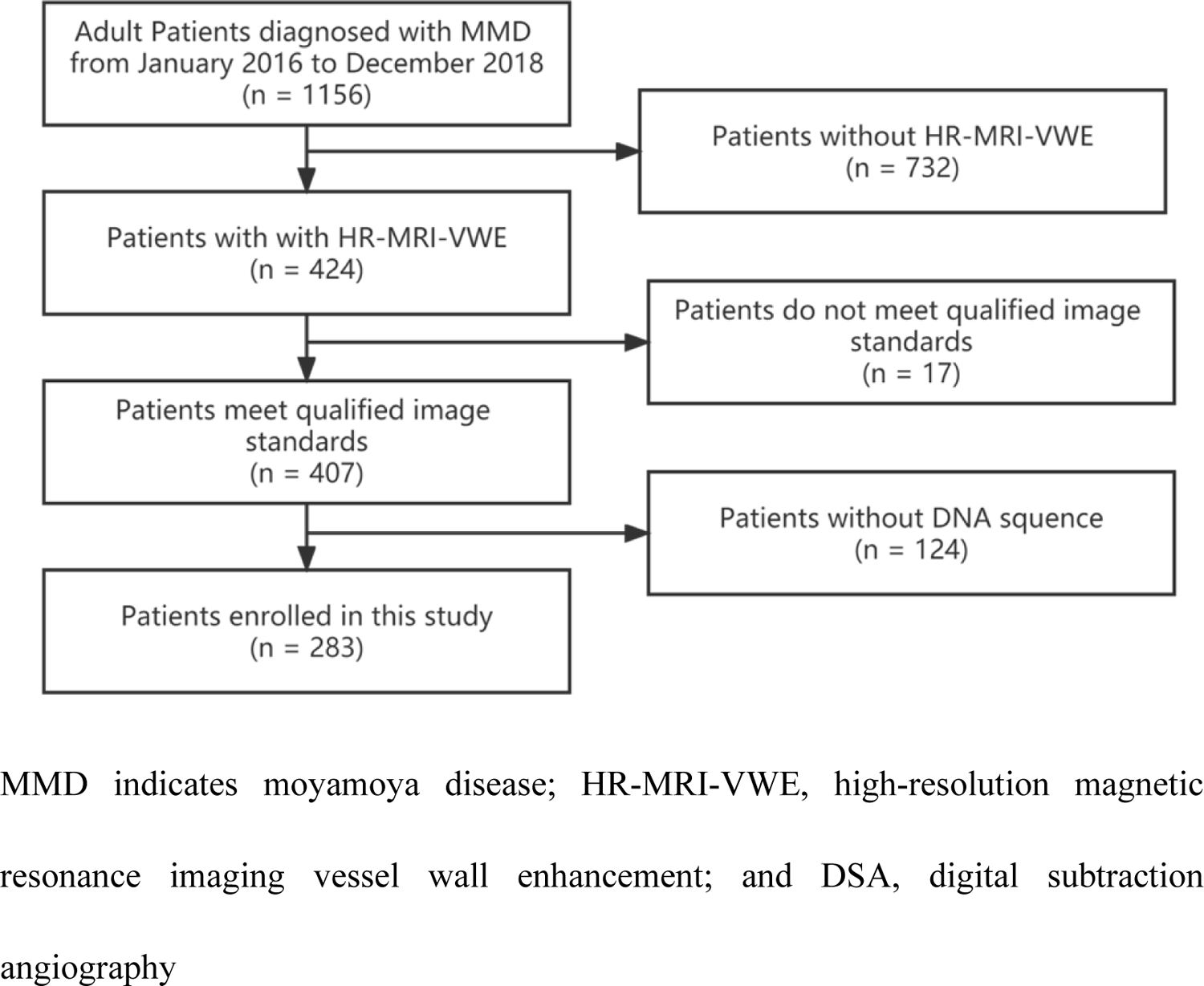
Flow chart of the study participants MMD indicates moyamoya disease; HR-MRI-VWE, high-resolution magnetic resonance imaging vessel wall enhancement; and DSA, digital subtraction angiography

**Figure 4.**
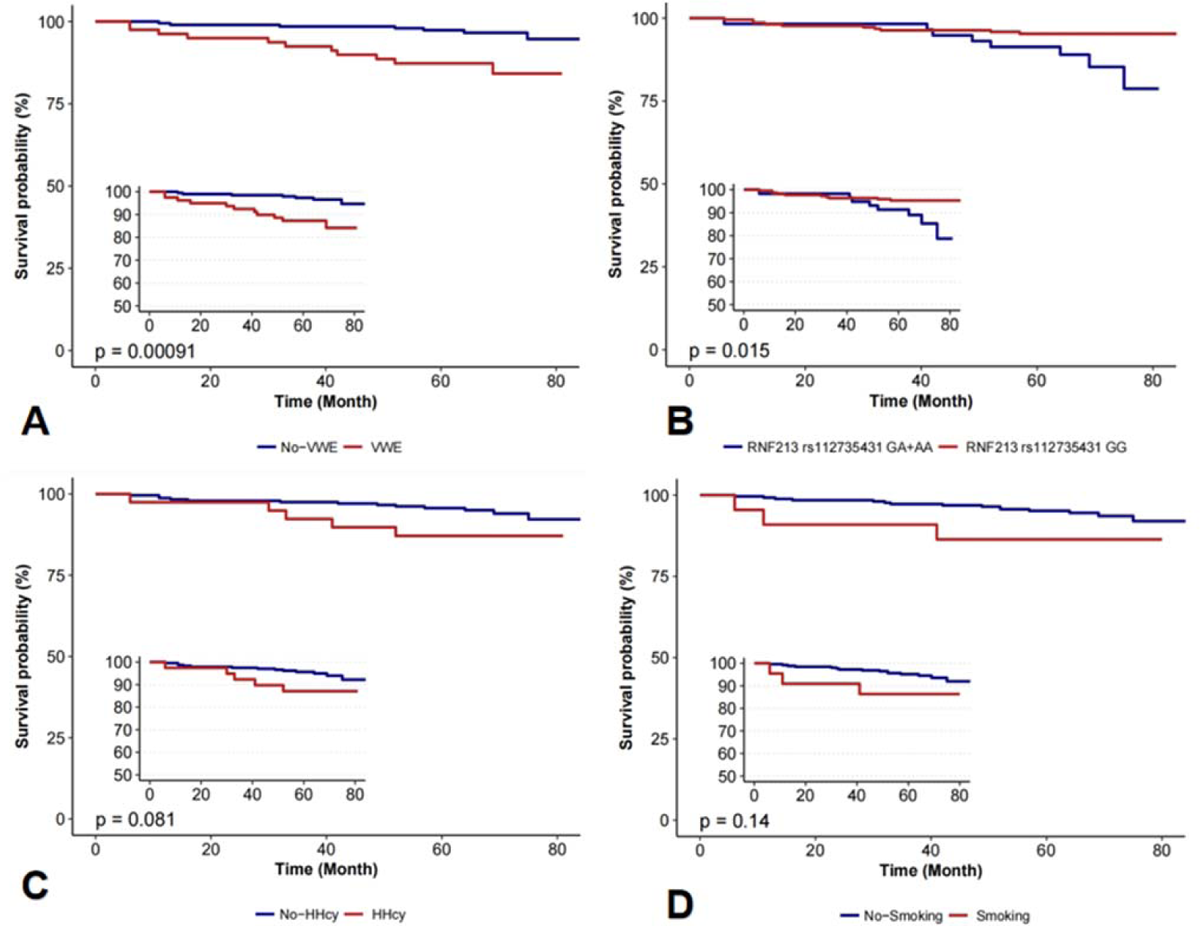
Kaplan–Meier analysis of different risk factors of recurrent stroke-free survival in patients with MMD.The risks of recurrent stroke identified by multivariate regression analysis were further analyzed using a log-rank test. Follow-up stroke-free survival was significantly different between VWE and non-VWE group (A) and between *RNF213* (RING finger protein 213) *p.R4810K* genotype GG and GA+AA(B). There was no significant difference for HHcy (C) or smoking (D).

**Table 1.**
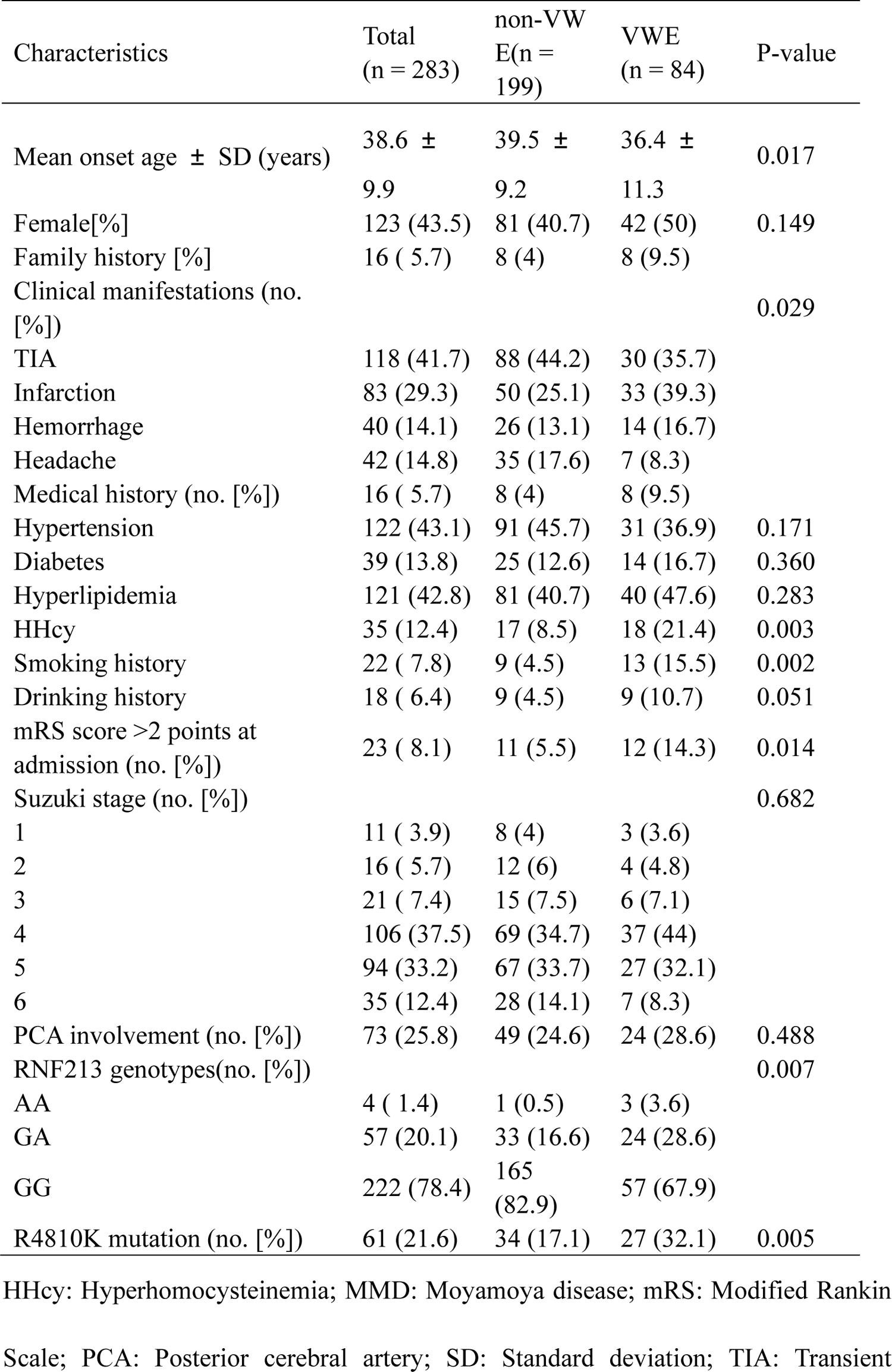
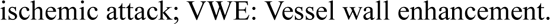
Clinical and genetic differences between VWE and non-VWE group

| Characteristics | Total<br>(n = 283) | non-VW<br>E(n =<br>199) | VWE<br>(n = 84) | P-value |
| --- | --- | --- | --- | --- |
| Mean onset age ± SD (years) | 38.6 ± 9.9 | 39.5 ± 9.2 | 36.4 ± 11.3 | 0.017 |
| Female[%] | 123 (43.5) | 81 (40.7) | 42 (50) | 0.149 |
| Family history [%] | 16 ( 5.7) | 8 (4) | 8 (9.5) |  |
| Clinical manifestations (no. [%]) |  |  |  | 0.029 |
| TIA | 118 (41.7) | 88 (44.2) | 30 (35.7) |  |
| Infarction | 83 (29.3) | 50 (25.1) | 33 (39.3) |  |
| Hemorrhage | 40 (14.1) | 26 (13.1) | 14 (16.7) |  |
| Headache | 42 (14.8) | 35 (17.6) | 7 (8.3) |  |
| Medical history (no. [%]) | 16 ( 5.7) | 8 (4) | 8 (9.5) |  |
| Hypertension | 122 (43.1) | 91 (45.7) | 31 (36.9) | 0.171 |
| Diabetes | 39 (13.8) | 25 (12.6) | 14 (16.7) | 0.360 |
| Hyperlipidemia | 121 (42.8) | 81 (40.7) | 40 (47.6) | 0.283 |
| HHcy | 35 (12.4) | 17 (8.5) | 18 (21.4) | 0.003 |
| Smoking history | 22 ( 7.8) | 9 (4.5) | 13 (15.5) | 0.002 |
| Drinking history | 18 ( 6.4) | 9 (4.5) | 9 (10.7) | 0.051 |
| mRS score >2 points at admission (no. [%]) | 23 ( 8.1) | 11 (5.5) | 12 (14.3) | 0.014 |
| Suzuki stage (no. [%]) |  |  |  | 0.682 |
| 1 | 11 ( 3.9) | 8 (4) | 3 (3.6) |  |
| 2 | 16 ( 5.7) | 12 (6) | 4 (4.8) |  |
| 3 | 21 ( 7.4) | 15 (7.5) | 6 (7.1) |  |
| 4 | 106 (37.5) | 69 (34.7) | 37 (44) |  |
| 5 | 94 (33.2) | 67 (33.7) | 27 (32.1) |  |
| 6 | 35 (12.4) | 28 (14.1) | 7 (8.3) |  |
| PCA involvement (no. [%]) | 73 (25.8) | 49 (24.6) | 24 (28.6) | 0.488 |
| RNF213 genotypes(no. [%]) |  |  |  | 0.007 |
| AA | 4 ( 1.4) | 1 (0.5) | 3 (3.6) |  |
| GA | 57 (20.1) | 33 (16.6) | 24 (28.6) |  |
| GG | 222 (78.4) | 165 (82.9) | 57 (67.9) |  |
| R4810K mutation (no. [%]) | 61 (21.6) | 34 (17.1) | 27 (32.1) | 0.005 |

HHcy: Hyperhomocysteinemia; MMD: Moyamoya disease; mRS: Modified Rankin
Scale; PCA: Posterior cerebral artery; SD: Standard deviation; TIA: Transient
ischemic attack; VWE: Vessel wall enhancement.

Of the 283 RNF213 p.R4810K sequenced patients, 222 (78.4%) were wild-type patients (G/G), 57 (20.1%) heterozygous patients (G/A), and 4 (1.4%) homozygotes (A/A). Compared with the patients in the non-VWE group, patients in the VWE group were included in more cases with G/A and A/A genotypes (P = 0.005). The *RNF213 p.R4810K* A allele carrying rate was 11.5%, and A allele frequencies were significantly higher in the VWE group than in the non-VWE group (P = 0.002). The SNP distribution of *p.R4810K* conformed to the Hardy-Weinberg. (Table 1)

### VWE-related risk factors and logistic regression results

Univariate logistic regression showed that age of onset, mRS score >2 points at admission, HHcy, Smoking history, and *RNF213* p.R4810K mutation were VWE risk factors for MMD (Table 2). Multivariate analysis showed that RNF213 p.R4810K mutation (OR, 2.30 [95% confidence interval (CI), 1.28-4.14], P = 0.006), mRS score >2 points at admission (OR, 3.02 [95% confidence interval (CI), 1.17-7.79], P = 0.022), HHcy (OR, 2.93 [95% CI, 1.32-6.48], P = 0.008), and history of smoking (OR, 3.52 [95% CI, 1.14-10.82], P = 0.028) were VWE risk factors after adjusting for gender, family history, hypertension, hyperlipidemia, diabetes, and alcohol drinking history.

**Table 2.** Logistic regression analyses of risk factors for VWE

| Variable | VWE |  |  |  |
| --- | --- | --- | --- | --- |
|  | Univariate |  | Multivariate* |  |
|  | OR (95% CI) | *P*-value | OR (95% CI) | *P*-value |
| Age of onset | 0.97 (0.94–0.99) | 0.019 | 0.98 (0.95–1.01) | 0.118 |
| Gender | 1.46 (0.87–2.43) | 0.15 | 1.21 (0.66–2.23) | 0.542 |
| mRS score >2 points at admission | 2.85 (1.2–6.75) | 0.017 | 3.02 (1.17–7.79) | 0.022 |
| Family history | 2.51 (0.91–6.94) | 0.075 | 1.59 (0.5–5.03) | 0.429 |
| Hypertension | 0.69 (0.41–1.17) | 0.172 | 0.64 (0.34–1.19) | 0.156 |
| Diabetes | 1.39 (0.68–2.83) | 0.362 | 2.05 (0.88–4.78) | 0.096 |
| Hyperlipidemia | 1.32 (0.79–2.21) | 0.283 | 1.46 (0.8–2.67) | 0.219 |
| HHcy | 2.92 (1.42–6) | 0.004 | 2.93 (1.32–6.48) | 0.008 |
| Smoking history | 3.87 (1.58–9.44) | 0.003 | 3.52 (1.14–10.82) | 0.028 |
| Alcohol drinking history | 2.53 (0.97–6.63) | 0.058 | 1.21 (0.34–4.29) | 0.769 |
| RNF213 p.R4810K mutation | 2.30 (1.28–4.14) | 0.006 | 2.29 (1.18–4.44) | 0.014 |

\*Multivariate regression analysis was adjusted for age, gender, hypertension, hyperlipidemia, diabetes, and alcohol drinking history.
CI: confidence interval; HHcy: hyperhomocysteinemia; OR: odds ratio; RNF213:
Ring finger protein 213; VWE: Vessel wall enhancement.

### VWE and follow-up incidences of stroke

Overall, 18 patients (6.4%) suffered a stroke during a median follow-up period of 63.9 ± 13.2 months (1.52% per person per year). Cox regression analysis demonstrated that the incidence rate of stroke was higher for patients with VWE (hazard ratio [HR]: 3.06; 95% CI: 1.03-9.1; *P* = 0.044) and the *RNF213* pR4810K variant (HR: 3.22; 95% CI: 1.13-9.19; *P* = 0.029) than in the non-VWE group (Table 3). K-M curves showed that stroke incidence was higher in the VWE group and in those with RNF213 pR4810K variants.

**Table 3.** Cox regression analyses of stroke-related risk factors of MMD during the follow-up period

| Variable | Follow-up stroke |  |  |  |
| --- | --- | --- | --- | --- |
|  | Univariate |  | Multivariate* |  |
|  | HR (95% CI) | P-value | HR (95% CI) | P-value |
| Age | 0.96 (0.92-1.01) | 0.086 | 0.98 (0.93-1.04) | 0.551 |
| Gender | 1.24 (0.49-3.15) | 0.653 | 0.88 (0.29-2.64) | 0.816 |
| VWE | 4.53 (1.75-11.75) | 0.002 | 3.06 (1.03-9.1) | 0.044 |
| RNF213 p.R4810K mutation | 4.45 (1.75-11.32) | 0.002 | 3.22 (1.13-9.19) | 0.029 |
| Hypertension | 0.93 (0.36-2.41) | 0.889 | 1.33 (0.44-3.98) | 0.610 |
| Diabetes | 0.79 (0.18-3.55) | 0.758 | 1.01 (0.21-4.98) | 0.987 |
| Hyperlipidemia | 1.1 (0.43-2.8) | 0.839 | 1.02 (0.36-2.9) | 0.969 |
| HHcy | 2.1 (0.69-6.4) | 0.19 | 1.84 (0.53-6.45) | 0.340 |
| Smoking history | 2.62 (0.75-9.13) | 0.131 | 1.63 (0.39-6.78) | 0.502 |
| Alcohol drinking history | 3.01 (0.86-10.48) | 0.083 | 1.98 (0.38-10.33) | 0.420 |

CI: confidence interval; HHcy: hyperhomocysteinemia; HR: hazard ratio; VWE: Vessel wall enhancement.

### Reproducibility

For detecting VWE, the intra-reader Kappa values were 0.93 and 0.88, and the inter-reader Kappa values were 0.85 and 0.83.

## Discussion

In this study, we compared the differences in clinical and genetic risk factors and long-term outcomes of MMD between the VWE group and the non-VWE group to further clarify the risk factors and possible causes of MMD progression. In the clinical aspect, early onset age, higher mRS score, presence of smoking history, and HHcy are confirmed in the VWE group than in the non-VWE group. In the genetic aspect, the *RNF213 pR4810K variant* is closely associated with VWE.

The MMD is a severe cerebrovascular disorder that causes the onset of strokes early. The etiology and pathophysiology of MMD remain unclear, and the prognosis varies.^20, 21^ VWE is a unique radiographic manifestation on HRMRI. Previous studies demonstrated that VWE in MMD is an important risk factor for rapid progress and poor prognosis.^13, 22^. In this study, we found that MMD patients with VWE had earlier onset age, a higher proportion of cerebral infarction and cerebral hemorrhage, higher mRS score, and higher stroke rate during the long-term follow-up, proving that VWE can be used as a predictor of poor prognosis in MMD.

Few studies have investigated correlations between gene polymorphisms and VWE in MMD patients. Notably, we identified a significant association between the *RNF213* pR4810K variant and VWE, and carriers of the A allele and GA+AA genotype were more susceptible to VWE. *RNF213* is the first susceptibility gene identified by both whole exon sequencing (WES) and genome-wide association analysis (GWAS) for MMD.^6-8^ *RNF213* plays diverse roles in angiogenic processes, including proliferation, migration, and capillary formation.^23^ As a means of imaging the vessel wall, VWE represents contrast agent accumulation in the vessel wall, and the mechanism may be functional disruption of the vascular endothelial barrier, leading to leakage of the contrast agent into the vessel wall^24^. The mechanism of VWE in MMD has not been clearly defined. Previous studies showed that the VWE phenomenon is associated with vascular endothelial damage^25–27^. Therefore, we hypothesize that the RNF213 p.R4810K variant may be responsible for damage to the vascular endothelium of the blood vessels, leading to the VWE phenomenon and MMD progression.

The Hcy level is a traditional vascular risk factor since high-level Hcy damages blood vessels by increasing oxidative stress and causing vascular endothelial dysfunction, angiogenesis disorders, and changes to the vascular intima structure.^28^ HHcy also has a mitogenic effect that promotes vascular smooth muscle proliferation^29, 30^. Previous research showed that HHcy is a risk factor for unilateral MMD, and it is associated with postoperative stroke in patients with MMD.^31^ However, the molecular mechanisms underlying HHcy in MMD remain unclear. We found higher serum Hcy levels in the VWE group than in the non-VWE group and an association between HHcy and VWE, suggesting that HHcy may participate in MMD progression. Therefore, controlling the Hcy level may be a potential non-surgical treatment for preventing stroke and progression in patients with MMD during preoperative and postoperative monitoring.

Smoking history increases the risk for ischemic and hemorrhagic stroke in people with MMD.^32, 33^ A previous study demonstrated that smoking affects the vascular endothelium by reducing nitrous oxide availability and promoting inflammation.^34^ Our data also provides evidence that smoking is a key risk factor for VWE in MMD. Therefore, like HHcy, we speculate that smoking may affect the cerebral vascular endothelium of MMD, leading to VWE and disease progression and increasing the incidence of stroke in the follow-up.

Our study had three limitations. First, we were unable to obtain pathological tissue of MMD in order to avoid harming the patient’s brain tissue, and the gene expression at the lesion site could not be measured. Second, we only evaluated whether the VWE was present in large intracranial arteries; the characteristics of the walls of perforating arteries, including tiny collateral circulation, could not be determined due to technical limitations. Third, the overarching goal of this study was to build upon previous research exploring the relationship between genotype, phenotype, VWE, and stroke, but our study design was unable to show causality. Future work will explore this relationship further.

In conclusion, VWE is a significant predictive factor of natural and postoperative stroke in patients with MMD. The *RNF213* pR4810K variant, HHcy and smoking are strongly associated with VWE in adults with MMD and may exacerbate MMD progression. Finally, HHcy and smoking are independent risk factors for VWE.; both are likely to induce the VWE phenomenon and consequently stroke by affecting vascular endothelial function or causing vascular endothelial damage. Controlling HHcy level and eliminating smoking may reduce vascular endothelial damage MMD.

## Data Availability

Owing to some sensitive information present in the study database, it is not freely available in the public domain currently. However, specific proposals for possible collaborations are welcome. Researchers interested in further information and collaboration are invited to contact the corresponding author Duan Lian via email

## Acknowledgments

No Funding

This study was supported by grants from the National Natural Science Foundation of China (grant numbers 82171280 and 82172021). Science and Technology Commission Project (2019-JCJQ-ZD-195-00).

## Disclosures

None

### Supplemental Material

STROBE guidelines checklis

## Non-standard Abbreviations and Acronyms

HRMRI: High-resolution magnetic resonance imaging

Hcy: Homocysteinemia

HHcy: Hyperhomocysteinemia

MRI: Magnetic resonance imaging

mRS: Modified Rankin Scale

MMD: Moyamoya disease

MMS: Moyamoya syndrome

PCA: Posterior cerebral artery

SNPs: Single nucleotide polymorphisms

VWE: Vessel wall enhancement

## Notes

### Competing Interest Statement

The authors have declared no competing interest.

### Author Declarations

The Research Ethics Committee of the Fifth Medical Center of the PLA General Hospital approved this study (reference number: 20150622).

## References

1. Scott RM, Smith ER. Moyamoya disease and moyamoya syndrome. The New England journal of medicine. 2009;360:1226–1237

2. Ihara M, Yamamoto Y, Hattori Y, Liu W, Kobayashi H, Ishiyama H, et al. Moyamoya disease: Diagnosis and interventions. The Lancet Neurology. 2022

3. Ya J, Zhou D, Ding J, Ding Y, Ji X, Yang Q, et al. High-resolution combined arterial spin labeling mr for identifying cerebral arterial stenosis induced by moyamoya disease or atherosclerosis. Ann Transl Med. 2020;8:87

4. Roder C, Hauser T-K, Ernemann U, Tatagiba M, Khan N, Bender B. Arterial wall contrast enhancement in progressive moyamoya disease. Journal of Neurosurgery. 2020;132:1845–1853

5. Dorschel KB, Wanebo JE. Genetic and proteomic contributions to the pathophysiology of moyamoya angiopathy and related vascular diseases. The application of clinical genetics. 2021;14:145–171

6. Liu W, Morito D, Takashima S, Mineharu Y, Kobayashi H, Hitomi T, et al. Identification of rnf213 as a susceptibility gene for moyamoya disease and its possible role in vascu lar development. PLoS One. 2011;6:e22542

7. Kamada F, Aoki Y, Narisawa A, Abe Y, Komatsuzaki S, Kikuchi A, et al. A genome-wide association study identifies rnf213 as the first moyamoya disease gene. Journal of Human Genetics. 2010;56:34–40

8. Miyatake S, Miyake N, Touho H, Nishimura-Tadaki A, Kondo Y, Okada I, et al. Homozygous c.14576g>a variant of rnf213 predicts early-onset and severe form of moyamoya disease. Neurology. 2012;78:803–810

9. Pathology RCot, Willis ToSOotCo, Health Labour Sciences Research Grant for Research on Measures for Infractable D. Guidelines for diagnosis and treatment of moyamoya disease (spontaneous occlusion of the circle of willis). Neurol Med Chir (Tokyo). 2012;52:245–266

10. von Elm E, Altman DG, Egger M, Pocock SJ, Gotzsche PC, Vandenbroucke JP, et al. The strengthening the reporting of observational studies in epidemiology (strobe) statement: Guidelines for reporting observational studies. Lancet. 2007;370:1453–1457

11. Zhang Y, Bao XY, Duan L, Yang WZ, Li DS, Zhang ZS, et al. Encephaloduroarteriosynangiosis for pediatric moyamoya disease: Long-term follow-up of 100 cases at a single center. J Neurosurg Pediatr. 2018:1–8

12. Hirano Y, Miyawaki S, Imai H, Hongo H, Ohara K, Dofuku S, et al. Association between the onset pattern of adult moyamoya disease and risk factors for stroke. Stroke. 2020;51:3124–3128

13. Lu M, Zhang H, Liu D, Liu X, Zhang L, Peng P, et al. Association of intracranial vessel wall enhancement and cerebral hemorrhage in moyamoya disease: A high-resolution magnetic resonance imaging study. J Neurol. 2021

14. Roy V, Ross JP, Pepin R, Cortez Ghio S, Brodeur A, Touzel Deschenes L, et al. Moyamoya disease susceptibility gene rnf213 regulates endothelial barrier function. Stroke. 2022:STROKEAHA120032691

15. Zhu B, Liu X, Zhen X, Li X, Wu M, Zhang Y, et al. Rnf213 gene polymorphism rs9916351 and rs8074015 significantly associated with moyamoya disease in chinese population. Ann Transl Med. 2020;8:851

16. Duan L, Wei L, Tian Y, Zhang Z, Hu P, Wei Q, et al. Novel susceptibility loci for moyamoya disease revealed by a genome-wide association study. Stroke. 2018;49:11–18

17. Liu S, Liu M, Li Q, Liu X, Wang Y, Mambiya M, et al. Association of single nucleotide polymorphisms of mthfr, tcn2, rnf213 with susceptibility to hypertension and blood pressure. Biosci Rep. 2019;39

18. Wang Y, Zhang Z, Wei L, Zhang Q, Zou Z, Yang L, et al. Predictive role of heterozygous p.R4810k of rnf213 in the phenotype of chinese moyamoya disease. Neurology. 2020

19. Yang Q, Zheng J, Chen W, Chen X, Wen D, Chen W, et al. Association between preadmission metformin use and outcomes in intensive care unit patients with sepsis and type 2 diabetes: A cohort study. Frontiers in medicine. 2021;8:640785

20. Hirano Y, Miyawaki S, Imai H, Hongo H, Teranishi Y, Dofuku S, et al. Differences in clinical features among different onset patterns in moyamoya disease. J Clin Med. 2021;10

21. Fox BM, Dorschel KB, Lawton MT, Wanebo JE. Pathophysiology of vascular stenosis and remodeling in moyamoya disease. Front Neurol. 2021;12:661578

22. Kathuveetil A, Sylaja PN, Senthilvelan S, Kesavadas C, Banerjee M, Jayanand Sudhir B. Vessel wall thickening and enhancement in high-resolution intracranial vessel wall imaging: A predictor of future ischemic events in moyamoya disease. AJNR Am J Neuroradiol. 2020;41:100–105

23. Roy V, Brodeur A, Touzel Deschenes L, Dupre N, Gros-Louis F. Rnf213 loss-of-function promotes angiogenesis of cerebral microvascular endothelial cells in a cellular state dependent manner. Cells. 2022;12

24. Millon A, Boussel L, Brevet M, Mathevet JL, Canet-Soulas E, Mory C, et al. Clinical and histological significance of gadolinium enhancement in carotid atherosclerotic plaque. Stroke. 2012;43:3023–3028

25. Perez FA, Oesch G, Amlie-Lefond CM. Mri vessel wall enhancement and other imaging biomarkers in pediatric focal cerebral arteriopathy-inflammatory subtype. Stroke. 2020;51:853–859

26. Larsen N, von der Brelie C, Trick D, Riedel CH, Lindner T, Madjidyar J, et al. Vessel wall enhancement in unruptured intracranial aneurysms: An indicator for higher risk of rupture? High-resolution mr imaging and correlated histologic findings. AJNR Am J Neuroradiol. 2018;39:1617–1621

27. Muraoka S, Taoka T, Kawai H, Okamoto S, Uda K, Naganawa S, et al. Changes in vessel wall enhancement related to the recent neurological symptoms in patients with moyamoya disease. Neurol Med Chir (Tokyo). 2021;61:515–520

28. Ganguly P, Alam SF. Role of homocysteine in the development of cardiovascular disease. Nutrition journal. 2015;14:6

29. Markan S, Sachdeva M, Sehrawat BS, Kumari S, Jain S, Khullar M. Mthfr 677 ct/mthfr 1298 cc genotypes are associated with increased risk of hypertension in indians. Molecular and Cellular Biochemistry. 2007;302:125–131

30. Meye C, Schumann J, Wagner A, Gross P. Effects of homocysteine on the levels of caveolin-1 and enos in caveolae of human coronary artery endothelial cells. Atherosclerosis. 2007;190:256–263

31. Ge P, Zhang Q, Ye X, Liu X, Deng X, Wang J, et al. Modifiable risk factors associated with moyamoya disease: A case-control study. Stroke. 2020;51:2472–2479

32. Yamagishi K, Iso H, Kitamura A, Sankai T, Tanigawa T, Naito Y, et al. Smoking raises the risk of total and ischemic strokes in hypertensive men. Hypertens Res. 2003;26:209–217

33. Boysen G, Nyboe J, Appleyard M, Sørensen PS, Boas J, Somnier F, et al. Stroke incidence and risk factors for stroke in copenhagen, denmark. Stroke. 1988;19:1345–1353

34. Yang L, Cheriyan J, Gutterman DD, Mayer RJ, Ament Z, Griffin JL, et al. Mechanisms of vascular dysfunction in copd and effects of a novel soluble epoxide hydrolase inhibitor in smokers. Chest. 2017;151:555–563

